# Assisted partner services for people who inject drugs: Index characteristics associated with untreated HIV in partners

**DOI:** 10.1101/2021.10.18.21265173

**Authors:** Ashley S. Tseng, Betsy Sambai, Aliza Monroe-Wise, Loice W. Mbogo, Natasha T. Ludwig-Barron, Sarah J. Masyuko, Bhavna H. Chohan, John D. Scott, William Sinkele, Joshua T. Herbeck, Carey Farquhar, Brandon L. Guthrie

## Abstract

**Objective:** Identify characteristics of persons who inject drugs living with HIV (PWID-LWH) associated with greater assisted partner services (APS) efficiency in identifying partners in need of HIV care and treatment services.

**Design:** Prospective cohort study

**Methods:** PWID-LWH (index participants) were enrolled and asked to provide contact information for sexual and injecting partners who were traced and offered HIV testing. APS efficiency was assessed by the number of indexes needed to interview (NNTI) to find one additional partner who was unaware of their HIV status or not on ART. We defined index participant characteristics associated with greater efficiency, defined as lower NNTIs.

**Results:** Among 783 indexes, the NNTI to identify one partner unaware of their HIV status was 7.1 and to identify one HIV-positive partner not on ART (regardless of status awareness) was 4.1. APS was provided to 977 partners and was more efficient in identifying partners who were not on ART (n=201) among indexes who were female (n=381, 49%; NNTI=2.9 vs. 5.7, p<0.001), unaware of their HIV status (n=74, 9.5%; NNTI=2.2 vs. 4.2, p=0.009), not on ART (n=158, 20%; NNTI=2.1 vs. 4.9; p<0.001), not enrolled in a methadone program (n=604, 77%; NNTI=3.3 vs. 10.4, p<0.001), reporting injecting <5 years (n=441, 56%; NNTI=3.3 vs. 5.0; p=0.005), or from Nairobi (n=452, 58%; NNTI=3.2 vs. 5.6, p<0.001).

**Conclusions:** Scaling up APS among PWID-LWH with certain characteristics could result in more efficient APS and greater partner engagement in HIV care.

## INTRODUCTION

A substantial barrier to ending the human immunodeficiency virus (HIV) epidemic is the large number of people living with HIV (PLWH) who remain undiagnosed and have not yet been linked to care. In 2019, there were approximately 38 million PLWH globally. It is estimated that 1 in 5 individuals living with HIV is unaware of their status, translating to 7 million PLWH who have not been tested[1]. In Kenya, the most recent national survey in 2020 found that 21% of adults living with HIV were unaware of their infection, a substantial decrease from 53% in 2013[2]. However, the proportion who are undiagnosed in key populations (KP), including people who inject drugs (PWID), was estimated to be higher[3], resulting in these groups likely contributing disproportionately to new infections annually.

HIV testing is a critical entry point for HIV prevention and treatment. Historically, emphasis in Kenya and other parts of sub-Saharan Africa has been on surveilling and testing the general population[4]. In recent years, significant progress has been made in scaling up HIV testing for KP[5]. In Kenya, an estimated 16,000 adults and young people inject drugs, with the vast majority concentrated in Nairobi, Mombasa, and Kisumu[1]. Despite relatively low absolute numbers, this population tends to be behaviorally vulnerable and their HIV prevalence was estimated to be 18%, compared to 5% among adults ages 15–49 in Kenya[1,5]. Linkage to HIV care is an important component of the care cascade and HIV status awareness is the first step, followed by continued engagement in HIV care.

The Kenyan Ministry of Health, committed to closing the HIV awareness gap and increasing engagement in care, has included the delivery of HIV assisted partner services (APS) as one of several strategies to increase uptake of HIV testing for the general population, and scaled up integrated out-of-facility initiatives through community outreach services to keep KP like PWID living with HIV (PWID-LWH) successfully engaged in care[6]. Voluntary APS, an evidence-based partner notification and testing strategy in which health practitioners collect contact information about the sexual or injecting partners of an index client with HIV, and then find partners, notify them of potential exposure, test them for HIV, and link them to care if living with HIV[7], is one approach to scale up HIV testing in a cost-effective way[8] and reduce barriers to disclosing HIV status to partners[9]. Studies of APS have shown the provision of partner services across a diverse range of communities and healthcare settings significantly and safely increases the uptake of HIV testing services and linkage to care for partners of persons with HIV[7,10–13].

Additional studies have examined the index participant characteristics associated with efficiency of HIV APS interventions, including geographical region, sex, and recency of diagnosis[14,15]. In Kenya, the effectiveness of APS in identifying new HIV diagnoses and linkage to care in the general population has been demonstrated [11,15,16]. However, APS has not been used specifically for HIV notification and linkage to care among PWID in Kenya.

The goal of this study was to identify characteristics of index participants (PWID-LWH) that were associated with identifying partners with HIV in need of engagement in care services (either newly diagnosed or already on ART). Knowing the specific index characteristics associated with higher APS efficiency can improve the design of public health intervention and surveillance programs to serve people affected by injection drug use.

## METHODS

### Study design and sites

This study is part of a prospective cohort study conducted in three counties in Kenya: Nairobi, Kilifi, and Mombasa. Kilifi and Mombasa, located on the eastern coast of Kenya, will be referred to as the ‘Coast’ region. Participant enrollment in the Study of HIV/Hepatitis C, APS, and Phylogenetics for Persons Who Inject Drugs (SHARP) began in March 2018 (U.S. NIH R01DA043409). Index participants were recruited from eight sites, including drop-in needle-syringe programs (NSP), medically assisted therapy (MAT) centers, and public health centers across these counties. Further details regarding study design and procedures can be found in the study protocol paper.[17]

### Study population and eligibility criteria

The study population was comprised of two groups: index participants (‘indexes’) and their sexual and/or injecting partner participants (‘partners’). Indexes are PWID-LWH who are accessing services at the study sites. To be included in the study, indexes had to have a positive HIV test result, be ≥18 years of age, injected drugs in the past year, able to provide written informed consent, and able to provide locator information for their sexual and/or injecting partners. Exclusion criteria included reporting sexual or physical intimate partner violence (IPV) within the last month. In addition to being identified by an index participant as a sexual and/or injecting partner, potential partner participants needed to be ≥18 years of age and able to provide informed consent. Both sexual and injecting partners were included in the definition of a ‘partner.’ Sexual partners were defined as people with whom indexes had sexual intercourse (i.e., vaginal, oral, or anal sex) with in the previous three years. Injecting partners were defined as anyone with whom indexes injected drugs with in the previous three years (regardless of whether they had shared needles).

### Study procedures

Trained health advisors administered structured questionnaires to collect information about demographics, sexual and injecting behaviors, drug use history, and HIV testing and care history via open data kit (ODK) software (ODK, 2017, Seattle, WA, USA). Indexes were asked to recall and provide contact and locator information for their sexual and/or injecting partners in the three years prior to enrollment. Community-embedded peer educators, who are former PWID with established relationships in the PWID communities that they serve, traced partners and referred them to study sites for HIV testing. APS services were provided regardless of study enrollment status. All indexes and all partners who tested positive for HIV or HCV at baseline completed a 6-month follow-up visit to assess linkage to and engagement in care.

This study received ethical approval from the University of Washington Human Subjects Division (IRB ID: STUDY00001536) and the Kenyatta National Hospital-University of Nairobi Ethics and Research Committee (IRB ID: KNH-ERC/R/195, P265/05/2017). All individuals provided written documentation of informed consent prior to participation.

### Analysis

#### Definition of Outcomes

The primary outcomes of interest were the number of partners identified through APS who were living with HIV and 1) unaware of their HIV status; or 2) not on ART at enrollment (both those newly diagnoses and those already aware of their positive status but not on ART). APS efficiency was assessed as the number of indexes that health advisors needed to interview (NNTI) to find one additional partner who was living with HIV and unaware of their HIV status or not on ART at enrollment. The NNTI was calculated as the total number of indexes interviewed, divided by the subsequent number of partners identified with the outcome of interest. Greater efficiency of APS applications is indicated by a lower NNTI value, thus a lower NNTI value is interpreted as the favorable outcome.

#### Statistical Analysis

Sociodemographic characteristics, stratified by sex, were compared descriptively for indexes and additionally described for partners. Participant sex, age, relationship status, enrollment region, housing status, awareness of HIV status at enrollment, current ART use, current enrollment in a methadone program, number of years injecting, shared needles history, and transactional sex history were categorical variables summarized using percentages. Poisson generalized linear models with an identity link were used to determine the efficiency of the APS intervention through calculated NNTI of indexes for the outcomes described above. We examined the association of the outcomes with index characteristics of interest by comparing the NNTI to find one additional partner with the outcome of interest between groups. Models were first assessed among the entire study population, then stratified by sex to evaluate potential effect modification of associations by sex. Univariate associations were explored (overall and for index men and women separately), then multivariate models were fit for index men and women separately for characteristics found to be statistically significant in the univariate analyses. ‘Engagement in transactional sex’ was defined as ever receiving money or other material gain for sex. A relationship status of ‘partnered’ was defined as an individual who cohabitates with someone (i.e., a live-in partner). Sexual or injecting partners were not differentiated in the analyses. Due to the nature of APS, partners could have been named multiple times by different indexes and enrolled in the study more than once, but the total number of partner study visits, not the number of unique partners, was used in this analysis to reflect this. Statistical tests were two-sided with a significance level of p<0.05.

## RESULTS

### Index Characteristics

A total of 783 index participants living with HIV were enrolled in the study as of February 2021, of whom 402 (51%) were men and 381 (49%) were women. The index women tended to be younger than the index men (median age 35 years [IQR: 30 to 40] vs. 38 years [IQR: 33 to 44]) (**Table 1**). Overall, a minority of indexes (30%) were married or partnered and most (86%) reported stable housing. The majority of men (56%) compared to only 30% of women reported injecting for 5 or more years. Most (81%) of the women and 25% of the men reported ever engaging in transactional sex. A small fraction of both men (4%) and women (9%) reported sharing needles in the past month, and 84% of index men and 76% of index women reported current ART use. Enrollment in a methadone program was reported by 28% men and 17% of women. The 783 indexes named 3664 partners overall (71% injecting partners, 18% sexual, and 12% dual sexual and injecting).

**Table 1.** Baseline characteristics of index participants living with HIV who identified sexual and injecting partners in the Study of HIV/Hepatitis C, APS, and Phylogenetics for Persons Who Inject Drugs (SHARP)

| Characteristic | Total<br>(N=783) | Men<br>(N=402) | Women<br>(N=381) |
| --- | --- | --- | --- |
|  | n (%) |  |  |
| Age (years) |  |  |  |
| 18-25 | 74 (9.5) | 25 (6.2) | 49 (12.9) |
| 26-35 | 274 (35.0) | 114 (28.4) | 160 (42.0) |
| 36-45 | 318 (40.6) | 185 (46.0) | 133 (34.9) |
| 46-67 | 117 (14.9) | 78 (19.4) | 39 (10.2) |
| Relationship status |  |  |  |
| Single | 287 (36.7) | 126 (31.3) | 161 (42.3) |
| Married or partnered | 237 (30.3) | 130 (32.3) | 107 (28.1) |
| Divorced or separated | 184 (23.5) | 104 (25.9) | 80 (21.0) |
| Widowed | 75 (9.6) | 42 (10.4) | 33 (8.7) |
| Enrollment region |  |  |  |
| Nairobi | 452 (57.7) | 189 (47.0) | 263 (69.0) |
| Coast | 331 (42.3) | 213 (53.0) | 118 (31.0) |
| Reported stable housing | 675 (86.2) | 339 (84.3) | 336 (88.2) |
| Aware of HIV status at enrollment | 709 (90.5) | 368 (91.5) | 341 (89.5) |
| Currently on antiretroviral therapy | 625 (79.8) | 336 (83.6) | 289 (75.9) |
| Currently enrolled in a methadone program | 176 (22.5) | 112 (27.9) | 64 (16.8) |
| Number of years injecting |  |  |  |
| <5 | 441 (56.3) | 175 (43.5) | 266 (69.8) |
| ≥5 | 337 (43.0) | 225 (56.0) | 112 (29.4) |
| Shared needles in past month | 47 (6.0) | 14 (3.5) | 33 (8.7) |
| Ever had transactional sex | 408 (52.1) | 101 (25.1) | 307 (80.6) |

### Partner Characteristics

APS was successfully provided to 3460 (94%) of the 3664 partners identified by the indexes, with a median of 4 partners tested per index, among both women and men. Among the partners, 65% were men and 35% were women. Partners were mostly (43%) ages 26-35 years. 20% of male partners were living with HIV compared to 43% of female partners. APS yielded 977 (28%) partners who were living with HIV, of whom 112 (11%) were unaware of their HIV status at enrollment and 89 (9%) were aware of their status but not on ART, for a total of 201 (21%) who were not on ART (regardless of status awareness). In the study population overall, 0.80 indexes needed to be interviewed to identify one partner living with HIV. When stratifying by sex, it was necessary to interview an average of 0.77 index women and 0.84 index men in order to identify one partner living with HIV (p=0.148).

### Index Characteristics Associated with Identifying Partners Unaware of Their Status

An average of 7.1 indexes were interviewed to identify one partner with HIV who was unaware of their status. Indexes who were unaware of their HIV status prior to study enrollment or not enrolled in a methadone program identified significantly more partners who were unaware of their HIV status (see Table, Supplemental Digital Content 1, which shows the NNTI to identify one additional partner unaware of their HIV status). An average of 5.9 index women were interviewed to identify one sexual or injecting partner who was unaware of their HIV status compared to 8.3 index men (p=0.078). The associations for identifying partners unaware of their status yielded similar results as the associations for identifying partners not on ART.

### Index Characteristics Associated with Identifying Partners Not Currently On ART

While most partners with HIV were already aware of their status, 201 (21%) of the partners with HIV were not on ART at the time of enrollment (including both those who were aware or unaware of their status at enrollment), requiring 4.1 indexes to be interviewed to identify one partner who was living with HIV and not on ART. Overall, the NNTI to identify a partner with HIV who was not on ART at study enrollment was 2.9 for index women versus 5.7 for index men (p<0.001). The NNTI to identify a partner not on ART was significantly lower in Nairobi compared to the Coast (p<0.001) (**Table 2**). The efficiency of finding partners living with HIV who were not on ART was greatest (i.e., lowest NNTI) among indexes who were unaware of their HIV status prior to enrollment (NNTI=2.2 vs. 4.2; p=0.009), those not currently on ART (NNTI=2.1 vs. 4.9; p<0.001), those not currently enrolled in a methadone program (NNTI=3.3 vs. 10.4; p<0.001), and those who reported injecting for less than 5 years (NNTI=3.3 vs. 5.0; p=0.005).

**Table 2.** Characteristics associated with the number of index participants needed to interview (NNTI) to identify one additional partner not on antiretroviral therapy at enrollment

| Index Characteristic |  | Total<br>(N=783) | Men<br>(N=402) | Women<br>(N=381) |
| --- | --- | --- | --- | --- |
|  |  | NNTI (p-value) |  |  |
| Sex | Men | 5.7 | - | - |
|  | Women | 2.9<br>(p<0.001*) | - | - |
| Age (years) | 18-25 | 3.4 | 6.3 | 2.7 |
|  | 26-35 | 3.4<br>(p=0.981) | 5.7<br>(p=0.862) | 2.6<br>(p=0.889) |
|  | 36-45 | 4.2<br>(p=0.373) | 5.4<br>(p=0.782) | 3.2<br>(p=0.551) |
|  | 46-67 | 5.1<br>(p=0.182) | 6.5<br>(p=0.946) | 3.5<br>(p=0.482) |
| Relationship status | Single | 3.5 | 6.6 | 2.6 |
|  | Married or<br>partnered | 4.8<br>(p=0.080) | 4.3<br>(p=0.142) | 5.6<br>(p=0.001*) |
|  | Divorced or<br>separated | 4.4<br>(p=0.253) | 7.4<br>(p=0.746) | 2.9<br>(p=0.670) |
|  | Widowed | 2.6<br>(p=0.183) | 6.0<br>(p=0.825) | 1.5<br>(p=0.061) |
| Enrollment region | Nairobi | 3.2 | 5.9 | 2.4 |
|  | Coast | 5.6<br>(p<0.001*) | 5.6<br>(p=0.827) | 5.6<br>(p<0.001*) |
| Reported stable housing | No | 4.2 | 9.0 | 2.4 |
|  | Yes | 3.9<br>(p=0.717) | 5.4<br>(p=0.120) | 3.0<br>(p=0.383) |
| Aware of HIV status at enrollment | No | 2.2 | 2.8 | 1.9 |
|  | Yes | 4.2<br>(p=0.009*) | 6.3<br>(p=0.060) | 3.1<br>(p=0.088) |
| Currently on antiretroviral therapy | No | 2.1 | 3.1 | 1.7 |
|  | Yes | 4.9<br>(p<0.001*) | 6.9<br>(p=0.017*) | 3.7<br>(p<0.001*) |
| Currently in a methadone program | No | 3.3 | 4.9 | 2.6 |
|  | Yes | 10.4<br>(p<0.001*) | 10.2<br>(p=0.008*) | 10.7<br>(p<0.001*) |
| Number of years injecting | <5 | 3.3 | 6.5 | 2.5 |
|  | ≥5 | 5.0<br>(p=0.005*) | 5.2<br>(p=0.376) | 4.7<br>(p=0.002*) |
| Shared needles in past month | No | 3.9 | 5.7 | 2.9 |
|  | Yes | 4.7<br>(p=0.519) | 7.0<br>(p=0.750) | 4.1<br>(p=0.241) |
| Ever had transactional sex | No | 5.2 | 6.7 | 2.7 |
|  | Yes | 3.2<br>(p<0.001*) | 4.0<br>(p=0.073) | 3.0<br>(p=0.737) |

\*Statistically significant at p<0.05

When stratifying by sex of the index, among both men and women, APS was more efficient in identifying partners not on ART for indexes who were not currently on ART (NNTI=3.1 vs. 6.9; p=0.017 for index men; NNTI=1.7 vs. 3.7; p<0.001 for index women) and were not enrolled in a methadone program (NNTI=4.9 vs 10.2; p=0.008 for index men; NNTI=2.6 vs. 10.7; p<0.001 for index women) (**Table 2**). The significant association for history of transactional sex and identifying partners not on ART was largely confounded by sex. Among index women, the efficiency of APS was greater among indexes in Nairobi compared to those on the Coast (NNTI=2.4 vs. 5.6; p<0.001). After adjusting for relationship status, the association between region and the number of partners who were not on ART remained significant (p<0.001). APS to find partners not on ART was also more efficient among index women who were single compared to those who were married or partnered (NNTI=2.6 vs. 5.6; p=0.001). The association was unchanged after adjusting for region (p=0.003). The efficiency of APS was greater for index women who injected for less than 5 years compared to those who had a longer history of injecting (NNTI=2.5 vs. 4.7; p=0.002); the association was non-significant after adjustment for region.

## DISCUSSION

In this study we successfully implemented APS for HIV exposure notification and testing among PWID in Kenya to identify injecting and sexual partners not currently on ART (either unaware of their positive HIV status or aware but not on ART). We found that APS was most efficient when it was conducted among index individuals who identify as women, those who were unaware of their HIV status prior to APS, those who were not currently on ART, those who were not currently enrolled in a methadone program, and those who had injected for less than 5 years. APS was also more efficient when conducted in Nairobi compared to coastal counties, but the higher efficiency in Nairobi was limited to APS conducted among index women. These findings demonstrate that scale-up of APS among PWID should consider index characteristics. This would ensure that resources committed to APS maximize the number of partners identified who would benefit from HIV testing and services to engage them in HIV care.

The efficiency of APS among PWID in Kenya compared favorably to its application in other populations and in other reviews [13,14,18,19]; the NNTI to identify a partner with HIV who was unaware of their status of 7.1 overall and 4.1 to identify a partner not on ART. In comparison, the NNTI to identify one new case of HIV ranges from 3 to 8 across general population studies in sub-Saharan Africa and Central Asia [13,14,18,19]. In sub-Saharan Africa, the NNTI to identify partners newly diagnosed with HIV in a study of Malawian pregnant women was 3.6 [20], in patients recruited from sexually transmitted infection clinics across Malawi was 4.5 [21], and in the Kenyan general population was 5.1 [13], which were all somewhat lower than what we observed in PWID-LWH in Kenya (7.1). The higher NNTI observed in our study reflects that most partners with HIV of PWID-LWH in our study already knew their HIV status (88%). We found that considerably fewer indexes needed to be interviewed to identify one partner with HIV not yet on ART (4.1). For PWID-LWH in Kenya, it may be most productive to focus APS on finding partners not on ART, regardless of their HIV status awareness, and ensuring that they are successfully engaged in care using targeted navigation services in order to promote engagement in HIV care. This differs from APS in many general populations where the focus is on identifying partners who are unaware of their HIV status who may not need the same level of support in accessing and being retained in care. The high number of partners who were aware their positive HIV status and not on ART indicates that existing HIV programs need to have services in place to successfully initiate and promote consistent ART use for partners with HIV who have not been engaged in care through previous testing programs.

APS in drop-in NSPs, MAT centers, and public health centers was efficient for both index men and women but was noticeably more efficient among index women compared to men. We also observed that the specific index characteristics associated with higher efficiency of finding partners with HIV who were unaware of their status or not on ART differed by the sex of the index individual. There was a non-significant trend of greater APS efficiency among index men who reported injecting for 5 or more years while the inverse trend was observed for index women: APS was significantly more efficient in those who injected for less than 5 years. Among index women, APS was more efficient among those who were single and from Nairobi, which was not observed among the index men. A prior study in Tanzania found that married partners were more likely to go to a health facility following HIV exposure notification using APS than unmarried partners, though they did not examine relationship status by female and male indexes separately or assess engagement in care outcomes [9]. Among the index men, those who reported ever engaging in transactional sex (25%) showed a non-significant trend of greater APS efficiency, which was not observed among the women who reported transactional sex (80%). This indicates that index men who report sexual behaviors that increase their vulnerability to HIV may provide an opportunity to reach partners who are less likely to be aware of their HIV status or on ART.

We found that APS was also more efficient for index men and women not currently on ART and not enrolled in methadone programs. Methadone programs, which administer methadone as an opioid medication to reduce withdrawal symptoms in people addicted to heroin, have been shown to reduce behavioral vulnerability to HIV [22,23]. PWID who are not enrolled in a methadone program can be considered to have increased vulnerability to HIV as these persons are likely actively injecting drugs. They in turn may have partners who are also more vulnerable to HIV and less likely to be successfully engaged in care if they know their status, leading to greater APS efficiency. This is consistent with our findings that the partners of those not in a methadone program were at higher risk of having undiagnosed or untreated HIV, arguing for greater investment in programs that reach PWID not enrolled in methadone programs. To better link PWID-LWH to both methadone and HIV care programs, PWID and their partners who present at HIV testing and counseling centers can be directed to drop-in centers where NSP and MAT services are offered and concurrently advised about ART initiation and continuation plans. Public health programs for PWID may achieve greater successes from APS by directing resources to fostering partnerships between methadone and HIV care programs as we move to latter steps in the care cascade.

This study is the first application of APS specific to a large population of PWID in sub-Saharan Africa to our knowledge, although APS has been successfully implemented among PWID in Central Asia[14] to identify both sexual and injecting partners of indexes. A limitation in our study was not including viral suppression data to examine factors associated with poor adherence or poor engagement in care among partners. However, we identified specific characteristics of indexes that can be used to inform future interventions in PWID. In resource-limited settings, it is important to focus HIV testing and initiation of ART efforts on PWID, a KP for HIV interventions. By focusing future efforts on PWID-LWH with these characteristics, we hope to help close the gap in HIV status awareness and to scale up engagement in care among partners with HIV.

In conclusion, APS was much more efficient in identifying sexual and injecting partners not on ART rather than partners unaware of their HIV status in our study. Future APS applications can more precisely engage PWID-LWH based on the index characteristics identified to be associated with greater APS efficiency in identifying partners with HIV not yet on ART.

## Data Availability

All data produced in the present study are available upon reasonable request to the authors.

## ACKNOWLEDGEMENTS

The authors would like to acknowledge the contributions of the SHARP participants, study team, community health workers, health advisors, clinical officers, and administrative staff.

## Contributors

J.T.H. and C.F. conceptualized the study. B.S. and B.L.G. managed the study data. A.S.T. conducted statistical analyses. A.S.T. drafted the article. All authors provided methodological and contextual input, reviewed, and approved the final article.

## REFERENCES

[1] UNAIDS, AIDSinfo, (2019). http://aidsinfo.unaids.org/ (accessed December 4, 2020).

[2] National AIDS and STI Control Programme (NASCOP), Preliminary KENPHIA 2018 Report, Nairobi, 2020.

[3] UNAIDS, Global HIV & AIDS Statistics — 2020 Fact Sheet, 2020. https://www.unaids.org/sites/default/files/media_asset/UNAIDS_FactSheet_en.pdf.

[4] J.M. Garcia-Calleja, National population based HIV prevalence surveys in sub-Saharan Africa: results and implications for HIV and AIDS estimates, Sex Transm Infect 82 (2006) iii64–iii70. https://doi.org/10.1136/sti.2006.019901.

[5] National AIDS Control Council of Kenya, Kenya AIDS Response Progress Report, 2014.

[6] National AIDS and STI Control Program (NASCOP), IMPROVING THE QUALITY AND EFFICIENCY OF HEALTH SERVICES IN KENYA: A Practical Handbook for HIV Managers and Service Providers on Differentiated Care, Nairobi, 2016.

[7] S. Dalal, C. Johnson, V. Fonner, C.E. Kennedy, N. Siegfried, C. Figueroa, et al., Improving HIV test uptake and case finding with assisted partner notification services, AIDS 31 (2017) 1867–1876. https://doi.org/10.1097/QAD.0000000000001555.

[8] M. Sharma, J.A. Smith, C. Farquhar, R. Ying, P. Cherutich, M. Golden, et al., Assisted partner notification services are cost-effective for decreasing HIV burden in western Kenya, AIDS 32 (2018) 233–241. https://doi.org/10.1097/QAD.0000000000001697.

[9] M. Plotkin, C. Kahabuka, A. Christensen, D. Ochola, M. Betron, M. Njozi, et al., Outcomes and Experiences of Men and Women with Partner Notification for HIV Testing in Tanzania: Results from a Mixed Method Study, AIDS Behav 22 (2018) 102–116. https://doi.org/10.1007/s10461-017-1936-x.

[10] C. Henley, G. Forgwei, T. Welty, M. Golden, A. Adimora, R. Shields, et al., Scale-Up and Case-Finding Effectiveness of an HIV Partner Services Program in Cameroon, Sex Transm Dis 40 (2013) 909–914. https://doi.org/10.1097/OLQ.0000000000000032.

[11] B.M. Wamuti, L.K. Erdman, P. Cherutich, M. Golden, M. Dunbar, D. Bukusi, et al., Assisted partner notification services to augment HIV testing and linkage to care in Kenya: study protocol for a cluster randomized trial, Implement Sci 10 (2015) 23. https://doi.org/10.1186/s13012-015-0212-6.

[12] C. Kahabuka, M. Plotkin, A. Christensen, C. Brown, M. Njozi, R. Kisendi, et al., Addressing the First 90: A Highly Effective Partner Notification Approach Reaches Previously Undiagnosed Sexual Partners in Tanzania, AIDS Behav 21 (2017) 2551– 2560. https://doi.org/10.1007/s10461-017-1750-5.

[13] P. Cherutich, M.R. Golden, B. Wamuti, B.A. Richardson, K.H. Ásbjörnsdóttir, F.A. Otieno, et al., Assisted partner services for HIV in Kenya: a cluster randomised controlled trial, Lancet HIV 4 (2017) e74–e82. https://doi.org/10.1016/S2352-3018(16)30214-4.

[14] K.M. Little, M. Kan, O. Samoylova, A. Rsaldinova, D. Saliev, F. Ishokov, et al., Implementation experiences and insights from the scaleJup of an HIV assisted partner notification intervention in Central Asia, J Int AIDS Soc 22 (2019). https://doi.org/10.1002/jia2.25313.

[15] S.J. Masyuko, P.K. Cherutich, M.G. Contesse, P.M. Maingi, B.M. Wamuti, P.M. Macharia, et al., Index participant characteristics and HIV assisted partner services efficacy in Kenya: results of a cluster randomized trial, J Int AIDS Soc 22 (2019). https://doi.org/10.1002/jia2.25305.

[16] P. Cherutich, M.R. Golden, B. Wamuti, B.A. Richardson, K.H. Ásbjörnsdóttir, F.A. Otieno, et al., Effectiveness of Partner Services for HIV in Kenya: A Cluster Randomized Trial, in: Conf. Retroviruses Opportunistic Infect., Boston, 2016.

[17] Monroe-Wise, L. Mbogo, B. Guthrie, D. Bukusi, B. Sambai, B. Chohan, et al., Peer-mediated HIV assisted partner services to identify and link to care HIV-positive and HCV-positive people who inject drugs: a cohort study protocol, BMJ Open 11 (2021) e041083. https://doi.org/10.1136/bmjopen-2020-041083.

[18] DiCarlo, A. Zerbe, Z.J. Peters, K. Frederix, J.P. Nkonyana, J.E. Mantell, et al., Use of Index Patients to Enable Home-Based Testing in Lesotho, JAIDS J Acquir Immune Defic Syndr 76 (2017) e61–e64. https://doi.org/10.1097/QAI.0000000000001486.

[19] P.M. Tih, F. Temgbait Chimoun, E. Mboh Khan, E. Nshom, W. Nambu, R. Shields, et al., Assisted HIV partner notification services in resourceJlimited settings: experiences and achievements from Cameroon, J Int AIDS Soc 22 (2019). https://doi.org/10.1002/jia2.25310.

[20] N.E. Rosenberg, T.K. Mtande, F. Saidi, C. Stanley, E. Jere, L. Paile, et al., Recruiting male partners for couple HIV testing and counselling in Malawi’s option B+ programme: an unblinded randomised controlled trial, Lancet HIV 2 (2015) e483–e491. https://doi.org/10.1016/S2352-3018(15)00182-4.

[21] L.B. Brown, W.C. Miller, G. Kamanga, N. Nyirenda, P. Mmodzi, A. Pettifor, et al., HIV Partner Notification Is Effective and Feasible in Sub-Saharan Africa: Opportunities for HIV Treatment and Prevention, JAIDS J Acquir Immune Defic Syndr 56 (2011) 437–442. https://doi.org/10.1097/QAI.0b013e318202bf7d.

[22] D. Otiashvili, G. Piralishvili, Z. Sikharulidze, G. Kamkamidze, S. Poole, G.E. Woody, Methadone and buprenorphine-naloxone are effective in reducing illicit buprenorphine and other opioid use, and reducing HIV risk behavior—Outcomes of a randomized trial, Drug Alcohol Depend 133 (2013) 376–382. https://doi.org/10.1016/j.drugalcdep.2013.06.024.

[23] S. Dvoriak, A. Karachevsky, S. Chhatre, R. Booth, D. Metzger, J. Schumacher, et al., Methadone maintenance for HIV positive and HIV negative patients in Kyiv: Acceptability and treatment response, Drug Alcohol Depend 137 (2014) 62–67. https://doi.org/10.1016/j.drugalcdep.2014.01.008.

