## Supplemental Table 1 for "Assisted partner services for people who inject drugs: Index characteristics associated with untreated HIV in partners"

**SUPPLEMENTAL DIGITAL CONTENT**

Supplemental Digital Content 1. The number of index participants needed to interview (NNTI) to identify one additional partner unaware of their HIV status

| Index Characteristic | | Total  (N=783) | Men  (N=402) | Women  (N=381) |
| --- | --- | --- | --- | --- |
|  |  | NNTI (p-value) | | |
| Sex | Men | 8.3 | - | - |
|  | Women | 5.9  (p=0.078) | - | - |
| Age (years) | 18-25 | 5.3 | 8.3 | 4.5 |
|  | 26-35 | 6.9  (p=0.415) | 8.6  (p=0.959) | 6.1  (p=0.416) |
|  | 36-45 | 7.5  (p=0.302) | 8.0  (p=0.946) | 6.9  (p=0.285) |
|  | 46-67 | 6.8  (p=0.490) | 8.6  (p=0.969) | 4.9  (p=0.845) |
| Relationship status | Single | 5.9 | 10.4 | 4.4 |
|  | Married or partnered | 7.1  (p=0.439) | 5.2  (p=0.069) | 10.6  (p=0.006*) |
|  | Divorced or separated | 8.8  (p=0.119) | 10.4  (p=0.997) | 7.3  (p=0.118) |
|  | Widowed | 7.5  (p=0.471) | 14.0  (p=0.621) | 4.7  (p=0.884) |
| Enrollment region | Nairobi | 7.0 | 11.0 | 6.9 |
|  | Coast | 6.8  (p=0.891) | 6.8  (p=0.103) | 5.6  (p=0.437) |
| Reported stable housing | No | 7.2 | 10.5 | 5.0 |
|  | Yes | 6.9  (p=0.875) | 8.0  (p=0.488) | 6.1  (p=0.615) |
| Aware of HIV status at enrollment | No | 3.7 | 4.9 | 3.1 |
|  | Yes | 7.6  (p=0.024*) | 8.9  (p=0.242) | 6.6  (p=0.059) |
| Currently on antiretroviral therapy | No | 5.6 | 7.3 | 4.8 |
|  | Yes | 7.4  (p=0.244) | 8.5  (p=0.701) | 6.4  (p=0.324) |
| Currently enrolled in methadone program | No | 6.1 | 7.2 | 5.3 |
|  | Yes | 13.3  (p<0.001*) | 13.6  (p=0.054) | 12.8  (p=0.010*) |
| Number of years injecting | <5 | 6.7 | 10.1 | 5.5 |
|  | $\geq$5 | 7.1  (p=0.738) | 7.2  (p=0.252) | 7.0  (p=0.379) |
| Shared needles in past month | No | 6.8 | - | 5.8 |
|  | Yes | 9.2  (p=0.450) | - | 6.4  (p=0.840) |
| Ever engaged in transactional sex | No | 9.0 | 10.6 | 5.6 |
|  | Yes | 5.7  (p=0.019*) | 5.1  (p=0.030*) | 6.0  (p=0.834) |

*****Statistically significant at p<0.05
